# Stem-like CD8+ T cells preserve HBV-specific responses in HBV/HIV co-infection

**DOI:** 10.1101/2025.03.30.25324898

**Authors:** Jay Preechanukul, Aljawharah Alrubayyi, Bo Sun, Edward Arbe-Barnes, Jonida Kokici, Frances Gorou, Sarun Prasitdumrong, Kelly A.S. da Costa, Natasha Fisher-Pearson, Noshin Hussain, Stephanie Kucykowicz, Indrajit Ghosh, Fiona Burns, Sabine Kinloch, Pedro Simoes, Sanjay Bhagani, Patrick T F Kennedy, Mala K Maini, Rachael Bashford-Rogers, Upkar S Gill, Dimitra Peppa

## Abstract

**Objective:** Chronic HBV infection disproportionately affects people living with HIV, who are often excluded from functional cure studies. This study investigates CD8^+^ T cell profiles in HBV mono-infection versus HBV/HIV co-infection, examining the impact of long-term therapy on virus-specific responses with the goal of informing therapeutic strategies for immune restoration.

**Design:** We analysed CD8^+^ T cell responses in 61 participants (HBV n=20, HBV/HIV n=20, HIV n=21), on suppressive antiviral therapy. We assessed transcriptomic and proteomic profiles, focusing on exhaustion markers alongside virus-specific functional capabilities.

**Results:** Transcriptomic analysis revealed a distinct signature in co-infection, with upregulation of genes associated with TCR signaling, inhibitory pathways and progenitor-exhausted markers (*XCL2, TCF7, PDCD1, IL7R*). This gene profile scored highly for a precursor exhausted (Tpex) CD8+ T cell signature, reflecting a "stemness" programme that maintains plasticity despite chronic antigen exposure. Proteomic analysis confirmed higher frequencies of precursor exhausted TCF-1^+^CD127^+^PD-1^+^ CD8^+^ T cells in co-infection, while HBV mono-infection showed predominance of terminally exhausted Tox^high^TCF-1^-^CD127^-^ cells. These differences correlated with more robust, polyfunctional HBV-specific responses in co-infection against surface and core antigens. Lower HBsAg levels and longer treatment duration in co-infection associated positively with Tpex populations and functional responses and inversely with terminal exhaustion.

**Conclusion:** Our findings demonstrate that individuals with well-controlled HBV/HIV co-infection maintain more robust CD8^+^ T cell responses with preserved stem-like properties supporting ongoing antiviral function. These results underscore the benefits of early antiretroviral intervention and the need for tailored immune-modulatory therapies to restore antiviral functionality in these diverse patient populations.

**WHAT IS ALREADY KNOWN ON THIS TOPIC:**

- Chronic hepatitis B virus (HBV) infection is marked by a progressive dysfunction of CD8⁺ T cells, which are crucial for antiviral responses. Traditionally these responses were thought to be more severely impacted in people with HBV/HIV co-infection.

**WHAT THIS STUDY ADDS:**

- Our study provides new insights into the heterogeneous functional profiles of HBV-specific CD8⁺ T cells in people with HBV and HBV/HIV co-infection in the current antiretroviral therapy (ART) era.
- People living with HBV/HIV co-infection suppressed on antivirals have a higher prevalence of precursor exhausted CD8⁺ T cells (Tpex), alongside more effective antiviral responses when compared to those with HBV mono-infection.
- Our data demonstrate intrinsic differences in T cell profiles, revealing a paradoxical increase in terminally exhausted CD8⁺ T cells in people with HBV mono-infection.

**HOW THIS STUDY MIGHT AFFECT RESEARCH, PRACTICE OR POLICY:**

- By providing a clearer understanding of CD8⁺ T cell dynamics in HBV mono-infection and HBV/HIV co-infection, our findings could inform the design of tailored immunotherapies aimed at revitalising antiviral responses.
- Furthermore, this research may influence practices regarding clinical management emphasising the need for early intervention strategies and individualised approaches tailored to T cell profiles rather than solely based on infection status.

## INTRODUCTION

Chronic hepatitis B (HBV) infection remains a significant health challenge, particularly among people living with HIV (PLWH), where approximately 10% are affected by concurrent chronic HBV ^1^. Yet people with co-infection are traditionally excluded from clinical trials, and their immune responses/HBV-specific immunity remain critically under-investigated, especially in the era of potent HBV-active antiretroviral therapies (Tenofovir Disoproxil Fumarate (TDF)/Tenofovir Alafenamide (TAF). These responses are important to elucidate in order to develop new and safe immunotherapeutic strategies and guide their involvement in the HBV-cure agenda.

The pathogenesis of chronic HBV infection is influenced by immune responses, particularly those mediated by CD8^+^ T cells, which are instrumental for viral control, and are thus promising targets for immunotherapeutic approaches aimed at achieving a functional cure (FC) [HBsAg loss] for chronic HBV (cHBV) infection^2^. cHBV results in diminished CD8^+^ T cell responses characterised by variable degrees of functional impairment, including upregulation of inhibitory molecules, specific transcriptional profiles and metabolic dysfunction, collectively reflecting a state of exhaustion due to prolonged antigen persistence^3–8^. HIV co-infection is thought to exacerbate these functional deficits, leading to more pronounced dysregulation of HBV-specific CD8^+^ T cell responses due to the combined influence of HIV and HBV^9^. In early studies, people with untreated co-infection had substantially fewer and narrower responses to HBV peptide stimulation compared to those with HBV mono-infection^10^. Although limited research examining responses to antiviral therapy prior to the introduction of highly active antiretroviral therapy (HAART) indicated a modest recovery of HBV immune responses, these responses remained suboptimal^11^. Notably, people with co-infection are more likely to achieve a FC following the introduction of ART^12^ ^13^; however, the effects of long-term viral suppression on reconstitution of HBV-specific responses remain unknown. Our recent findings have indicated that, in the context of long-term therapy, people with HBV/HIV co-infection exhibit more favourable immunological profiles, evidenced by better preserved natural killer (NK) cell responses and lower levels of HBsAg and peripheral surrogates of HBV activity compared to those with HBV mono-infection^14^. Given the critical role of CD8+ T cells in HBV control, there is an urgent need for increased insights into the specific profiles of CD8+ T cells in people with HBV/HIV.

Recent studies have revealed that exhausted CD8^+^ T cell populations are not homogeneous; they exhibit developmental and functional diversity, including the presence of stem-like progenitors expressing T cell factor 1 (TCF-1)^15^. These progenitors or precursors of exhausted CD8^+^ T cells (Tpex) retain the potential to mount effective immune responses^16^, allowing for the possibility of therapeutic intervention aimed at restoring immune efficacy^17^.

Importantly, the nature of T cell dysfunction in cHBV may differ from other chronic infections. Emerging evidence suggests that T cell dysfunction in cHBV infection incorporates traits more reminiscent of a state of tolerance or anergy^18^, underscoring the need for a more nuanced understanding of these dynamics, particularly in the under-explored context of co-infection.

Understanding the heterogeneity of HBV-specific CD8⁺ T cells in various populations is therefore an important step for identifying potential targets for immunotherapy and uncovering novel molecular pathways that could enhance antiviral immune responses.

In this study, we aimed to address these critical gaps in understanding the immune responses of individuals with HBV/HIV co-infection compared to those with HBV mono-infection in the current ART era. We hypothesised that earlier initiation and long-term viral suppression with potent antiretroviral therapy might preserve functional CD8+ T cell responses in co-infection, contrary to traditional expectations of more severe immune dysfunction. We performed a detailed analysis at the single-cell level, complemented by assessments of functional virus-specific responses, to enhance our knowledge of the immunological landscape in these patient populations.

## MATERIALS AND METHODS

### Patient populations

A total of n=20 patients with HBV/HIV, n=20 people with HBV and n=21 with HIV were recruited at Mortimer Market Centre for Sexual Health and HIV research, the Ian Charleson Day Centre at the Royal Free Hospital (London, UK) or The Royal London Hospital (London, UK) following written informed consent as part of a study approved by the local ethics committee (Berkshire (REC 16/SC/0265) and London Bridge (REC 17/LO/0266) and conformed to the Helsinki declaration principles. All participants were negative for HCV infection and confirmed HCMV seropositive. PBMCs, plasma and serum were collected as part of this study. Additional demographic and clinical information can be found in **supplementary table 1**.

### Single cell Sequencing

#### Library preparation and sequencing

PBMCs with >96% viability were used^14^. Library construction was performed using the 10x next GEM Chips Chromium single cell library 5’ construction kit (10x Genomics, Pleasanton) as per the manufacturer’s protocol. Briefly, a maximum of 3000 cells were combined with barcoded single cell VDJ gel beads, cells and partitioning oil onto Chromium Next GEM Chip K. The resulting 10x Barcoded, full-length cDNA was recovered and amplified overnight via PCR with primers against common 5’ and 3’ ends added during the GEM reverse transcription process. Resulting cDNA was purified using SPRIselect beads (Beckman Coulter, High Wycombe, UK). The amount and quality of cDNA was determined using TapeStation (Agilent, UK). Purified libraries were analyzed by Novogene. Donor characteristics analysed by single-cell RNAseq data are described in **supplementary table 2.**

### Single-Cell RNA Sequencing Preprocessing and Procedures

Single-cell RNA sequencing (scRNA-seq) preprocessing and downstream analyses for this study have been previously described in detail^14^. Briefly, we excluded any cell barcodes with fewer than 200 detected transcripts or more than 10% mitochondrial-encoded genes, retaining only cells expressing fewer than 3000 genes. We considered genes if they were expressed in at least five cells. After confirming that our data and the Azimuth PBMC reference atlas^19^ showed similar distributions of transcripts and gene counts, we demultiplexed hash-tagged samples using the Scanpy external implementation of Hashsolo^20^ and excluded doublets using Solo. For dataset integration, raw counts from the top 3000 highly variable genes were used to train a scVI model, which generated 20 latent dimensions for embedding. We then computed a nearest neighbor graph on these dimensions for cell clustering via iterative Leiden clustering at increasing resolutions to capture both coarse and fine cell type heterogeneity. Automated cell type annotations were performed with Cell typist ^21^, followed by manual validation based on marker gene expression and hierarchical clustering.

Differential gene expression was calculated with Seurat’s FindMarkers function (using DESeq2 as the statistical backend), controlling for variable sample sizes across study groups. Functional enrichment analysis was carried out using the XGR package for pathway-level assessment and GSEApy for gene set enrichment analysis^22^. Differential abundance analysis was performed with the R implementation of Milo^23^. For the Milo buildGraph function, k was set to 40, and all 128 dimensions of the DRVI latent embedding were used. Where described, gene set scores were calculated with Scanpy’s score_genes function. The Tpex-score genes were derived from Zheng et al.^24^. For the unbiased gene program discovery analysis, DRVI was implemented with default parameters^25^.

### *Ex-vivo* phenotypic analysis of CD8^+^ T cells

The fluorochrome-conjugated antibodies utilised are detailed in **supplementary table 3.** Briefly, cryopreserved PBMCs were thawed and allowed to rest at 37 °C in a complete RPMI medium (Penicillin-Streptomycin, L-Glutamine, HEPES, non-essential amino acids, 2-Mercaptoethanol, and 10% Fetal Bovine Serum (FBS)). The cells were washed, resuspended in PBS, and incubated for 20 minutes at 4°C with various antibody combinations along with a fixable live/dead stain (Invitrogen). The cells were subsequently fixed and permeabilised to facilitate the detection of intracellular antigens. For the identification of intranuclear markers, the Foxp3 intranuclear staining buffer kit (eBioscience) was used according to the manufacturer’s protocol. Total PBMCs were stained with APC-labeled HBV (core 18-27, envelope 183–191, envelope 335–343, and envelope 348–357), HCMV (pp65 495-504), and HIV (Gag 77-86) dextramers (Immudex) at 37°C for 15 minutes in complete RPMI with 10% FBS. After pelleting, cells were further stained as above. Dextramer staining was deemed positive if a distinct population (>0.02%) was identified. Data acquisition was conducted on a BD Fortessa X20 via BD FACSDiva8.0 (BD Bioscience), and the resulting data analysis was carried out using FlowJo 10 (TreeStar). The gating strategy for identifying CD8^+^ T cells is illustrated in **supplementary figure 2A**. Stochastic neighbor embedding (SNE) analysis was performed on the MRC Cytobank platform to visualise high-dimensional data.

### Functional assessment of virus-specific CD8^+^ T cells

Intracellular cytokine staining (ICS) was performed as previously described^26^. PBMCs were thawed and allowed to rest at 37 °C in complete RPMI medium with 5% CO_2_. PBMCs were then stimulated for 12-16 hours using 3 μg/mL of overlapping peptide pools from HBV (large envelope and capsid), HIV-1 Gag, or positive control peptide pools derived from cytomegalovirus (HCMV), Epstein Barr virus (EBV) and influenza virus (CEF pool) **(supplementary table 4)**, alongside 0.005% DMSO as a negative control. This was done in the presence of αCD28/αCD49d co-stimulatory antibodies (1 μg/ml), Brefeldin A (eBioscience). Post-stimulation, cells were stained with anti-CCR7 at 37 °C for 30 minutes, then surface stained at 4 °C for 20 minutes along with a live/dead stain. Following fixation and permeabilisation, intracellular cytokines (IFN-γ APC, CD154 PE-Cy7, TNF-α FITC, IL-2 PerCP-eFluor 710) were assessed. Data acquisition was performed using a BD Fortessa X20, and analysis was conducted using FlowJo 10. Virus-specific CD8+ T cells were identified as those expressing combinations of IFN-γ, TNF-α, IL-2. A detailed list of antibodies used is found in **supplementary table 3.**

### Statistical methods

Prism 9 (Graphpad, San Diego) and R (URL https://www.R-project.org/)was used for statistical analysis. The Mann–Whitney U-test was used for single comparisons of groups, and the Wilcoxon-paired t-test was used to compare two paired groups. Correlation analysis was carried out with non-parametric Spearman correlation. Kruskal Wallis testing followed by Dunn test was used for pairwise comparison between gene module scores. The statistical significance is indicated in the figures. (\**p* < 0.05, \*\**p* < 0.01, \*\*\**p* < 0.001, and \*\*\*\**p* < 0.0001).

## RESULTS

### Single cell analysis of CD8^+^ T cells shows reduced cytotoxicity gene expression and enriched TCR signalling pathways in HBV/HIV co-infection

To explore the effects of HBV and combined effect of HBV/HIV on CD8^+^ T cell populations, we utilised available single-cell sequencing PBMC data (using 10X Chromium (5’ transcriptome) from n = 5 HBV/HIV and n = 6 HBV donors, predominately on TDF treatment **(supplementary table 2)**^14^. To account for the effect of human cytomegalovirus (HCMV), all donors were confirmed human HCMV-seropositive. CD8^+^ T cell subsets – naive CD8^+^ T cell (TN), central memory (TCM) CD8^+^ T cell, effector memory (TEM) CD8^+^ T cell, terminally differentiated effector memory (TEMRA) CD8^+^ T cell and memory precursor effector cell (MPECS), were identified by clustering based on their differentially expressed genes (DEGs) and visualised on a uniform manifold approximation and projection (UMAP) **(figure 1A-B)**.

**Figure 1.**
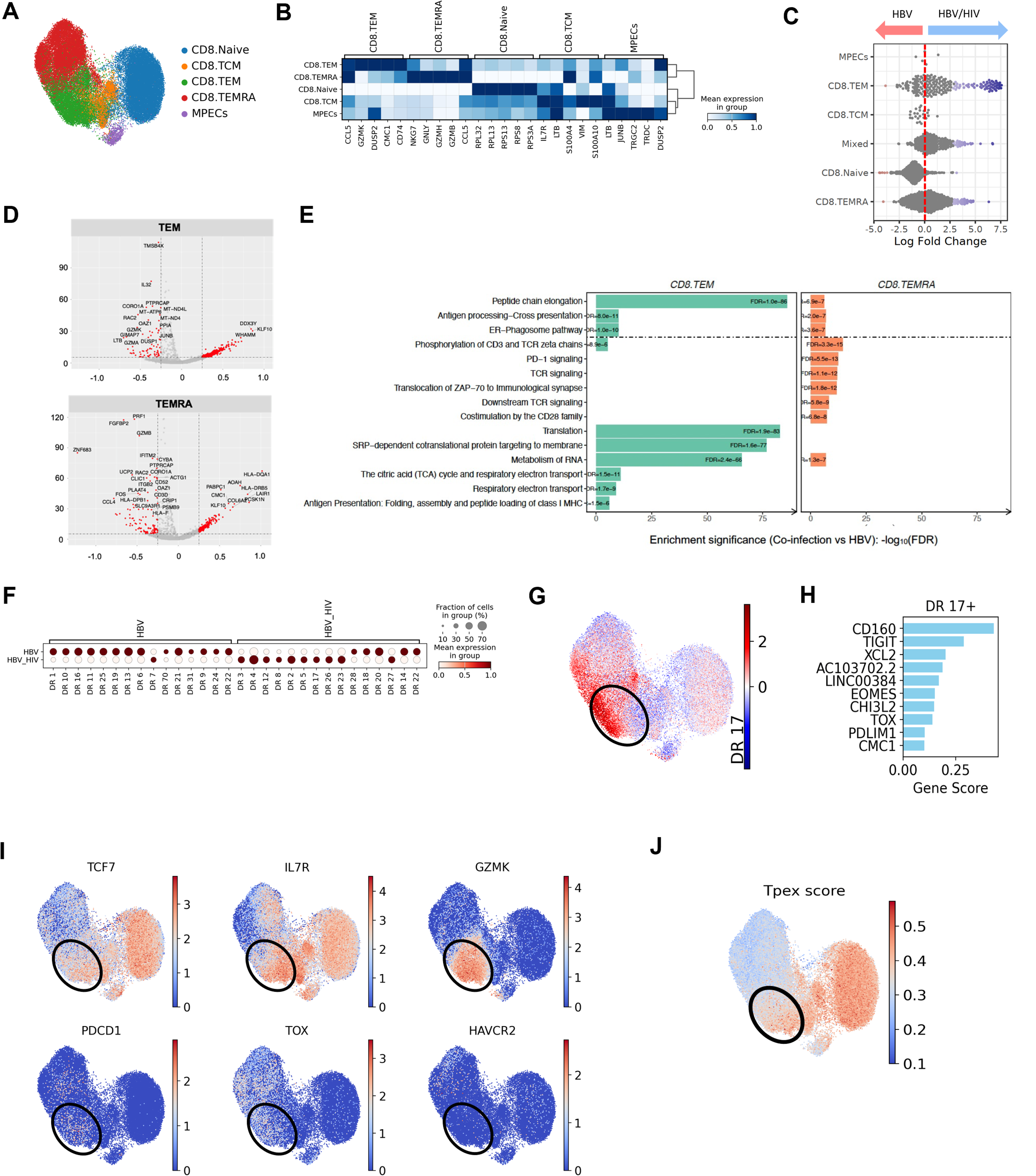
Transcriptomic profiles of CD8^+^ T cell subsets in HBV mono-infection vs. HBV/HIV co-infection. (A) UMAP embedding of n = 51,843 CD8^+^ T cells from n = 11 individuals (HBV n=6; HBV/ HIV co-infection, n=5). (B) Expression heatmap showing distinct gene expression profiles for CD8^+^ T cell subsets. Genes were selected as the 5 most significant differentially expressed genes per cluster. (C) Milo cell neighbourhood differential abundance plots of the significantly enriched neighbourhoods between HBV mono-infection vs. HBV/HIV co-infection (FDR <0.05). (D) Volcano plot of DESeq2 differential gene expression analysis; significant genes (FDR <0.05 and average log2FC >/< 0.25) are highlighted. The direction is in favour of HBV/HIV co-infection, for example, positive fold changes are upregulated in HBV/HIV co-infection versus HBV mono-infection. (E) Bar plot of most significantly enriched (FDR <0.005) reactome pathways in HBV/HIV co-infection versus HBV mono-infection. (F) Differential expression analysis of DRVI derived gene programmes (denoted numerically as DRs). (G) UMAP embedding showing activity of DR17. (H) Top identified genes for DR17. (I) The expression of relevant genes on UMAP. (J) The activity of the Tpex score on UMAP. Abbreviations: HBV, Hepatitis B Virus; HIV, Human immunodeficiency virus; FDR, false discovery rate; UMAP, uniform manifold approximation and projection; DR, Dimensionality reduction.

To gain insight into cellular differential abundance and compositional shifts between HBV mono-infection vs. HBV/HIV co-infection, MiloR was used for differential abundance testing. While naive CD8⁺ T-cells trended towards more abundance in HBV mono-infection, we found that most of the significant changes driven by HBV/HIV co-infection were within the CD8⁺ TEM compartment **(figure 1C)**. Comparing the transcriptional profiles of CD8⁺ TEM cells between patient groups, donors with co-infection showed an up-regulation of *KLF10*, which encodes an effector protein of transforming growth factor beta (TGF-β) signaling ^27^, *DDX3Y*, which is involved in RNA binding and the formation of intramolecular interactions, and *WHAMM*, which encodes a protein that mediates membrane dynamics and cytoskeletal organisation within cells^28^ **(figure 1D)**. In contrast, several cytotoxicity genes were upregulated in the CD8^+^ TEM and TEMRA clusters of mono-infected patients (**figure 1D**), correlating with higher cytotoxicity scores observed in these populations in HBV mono-infection **(supplementary figure 1A)**. Genes driving CD8⁺ T cell remodeling were reflected in differentially regulated gene pathways, with enrichment of TCR downstream signaling, co-stimulation, PD-1 signalling and TCA cycle and translation observed in the CD8^+^ T cell populations in the co-infection group **(figure 1E**).

Next, we sought to identify de novo gene programs that might be differentially used across mono-infection and co-infection. To do so, we used DRVI ^25^ (an unsupervised deep generative model that learns nonlinear, disentangled representations of single-cell omics data, allowing us to discover biologically meaningful latent dimensions and nonlinear gene programs. Applying DRVI led to the identification of 39 gene programs **(supplementary figure 1B–C)**. Notably, “DR17” was differentially upregulated in the same neighborhood that MiloR analysis pinpointed as being enriched in HBV/HIV co-infection **(figure 1F–G)**.

Among the relevant genes in DR17 were *CD160, TIGIT, XCL2*, *CHI3L2*, *EOMES*, *TOX*, *PDLIM1*, and *CMC1* **(figure 1H)**. Upregulation of *CHI3L2* (involved in inflammation regulation) ^29^ and *CMC1* (involved in mitochondrial function)^30^ suggest cellular and metabolic adaptations of CD8⁺ T cells in co-infection. Of particular interest, *XCL2* has been associated with progenitor or precursors of exhausted (Tpex) T cells^31^. Further interrogation showed that T_pex_ markers (*TCF7*, *PDCD1*, *IL7R*, *GZMK*, *TOX*) were present and *HAVCR2* (which encodes TIM-3, characterising terminally exhausted populations) was notably absent in this neighbourhood **(figure 1I)**, consistent with a Tpex phenotype, associated with a favourable response to combination therapy in hepatocellular carcinoma^31^. In addition, co-expression of inhibitory receptors (*CD160*, *TIGIT*) and exhaustion-associated transcription factors (*EOMES*, *TOX*) supports a Tpex-like transcriptional signature in these cells^17^ ^32^. To validate whether this co-infection-enriched neighbourhood indeed displayed Tpex-like features, we calculated a Tpex score based on Tpex-associated genes identified through a pan-cancer single-cell atlas^24^. As expected, this region scored highly for the Tpex signature alongside naïve and MPEC populations, likely reflecting a shared “stemness” gene programme that underlies both memory-precursor and stem-like exhausted states **(figure 1J)**. Overall, these signatures suggest an adapted immune state in HBV/HIV co-infection where CD8⁺ T cells counter-balance inhibitory mechanisms with stemness features that could maintain plasticity and functional potential despite chronic antigen exposure.

### Precursors of exhausted CD8^+^ T cells are enriched in HBV/HIV co-infection

To explore whether the proteomic profile of CD8^+^ T cells in individuals with HBV mono-infection (n = 20) and HBV/HIV co-infection (n = 20) mirrors transcriptomic signatures, we conducted a phenotypic analysis within our wider cohort. Matched donors with HIV mono-infection (n = 21) virally suppressed on antiretroviral therapy were utilised as a comparator group (**supplementary table 1)**. Using a curated cytometry panel and gating strategy **(supplementary figure 2A)**, we evaluated CD8^+^ T cell subsets. As expected, higher frequencies of CD8+ T cells were observed in the context of HIV infection (**supplementary figure 2B**). HBV/HIV co-infection and HIV mono-infection showed increased frequencies of CD8^+^ TEM and TEMRA, while HBV mono-infection predominantly exhibited CD8^+^ naive T cells, consistent with findings from single-cell analyses (**supplementary figure 2C-D**).

Using an unbiased global t-distributed stochastic neighbour embedding (t-SNE) high dimensional analysis and FlowSOM clustering, we further identified a distinct subpopulation (metacluster) that was more prominent in people with HBV/HIV co-infection relative to the other groups (**figure 2A**). This metacluster 1, which was more abundant in HBV/HIV co-infection, displayed a phenotypic signature characterised by the presence of CD45RA^-^CCR7^+/-^TIM3^-^PD1^+/-^CD127^high^TCF1^+^ **(figure 2A-B**). Manual gating analysis further confirmed increased frequencies of CD127^high^TCF1^+^ CM/TEM CD8+ T cells, which were particularly expanded within specific donors with HBV/HIV co-infection **(figure 2B-C)**, suggesting a less differentiated memory-like state with preserved stemness properties.

**Figure 2.**
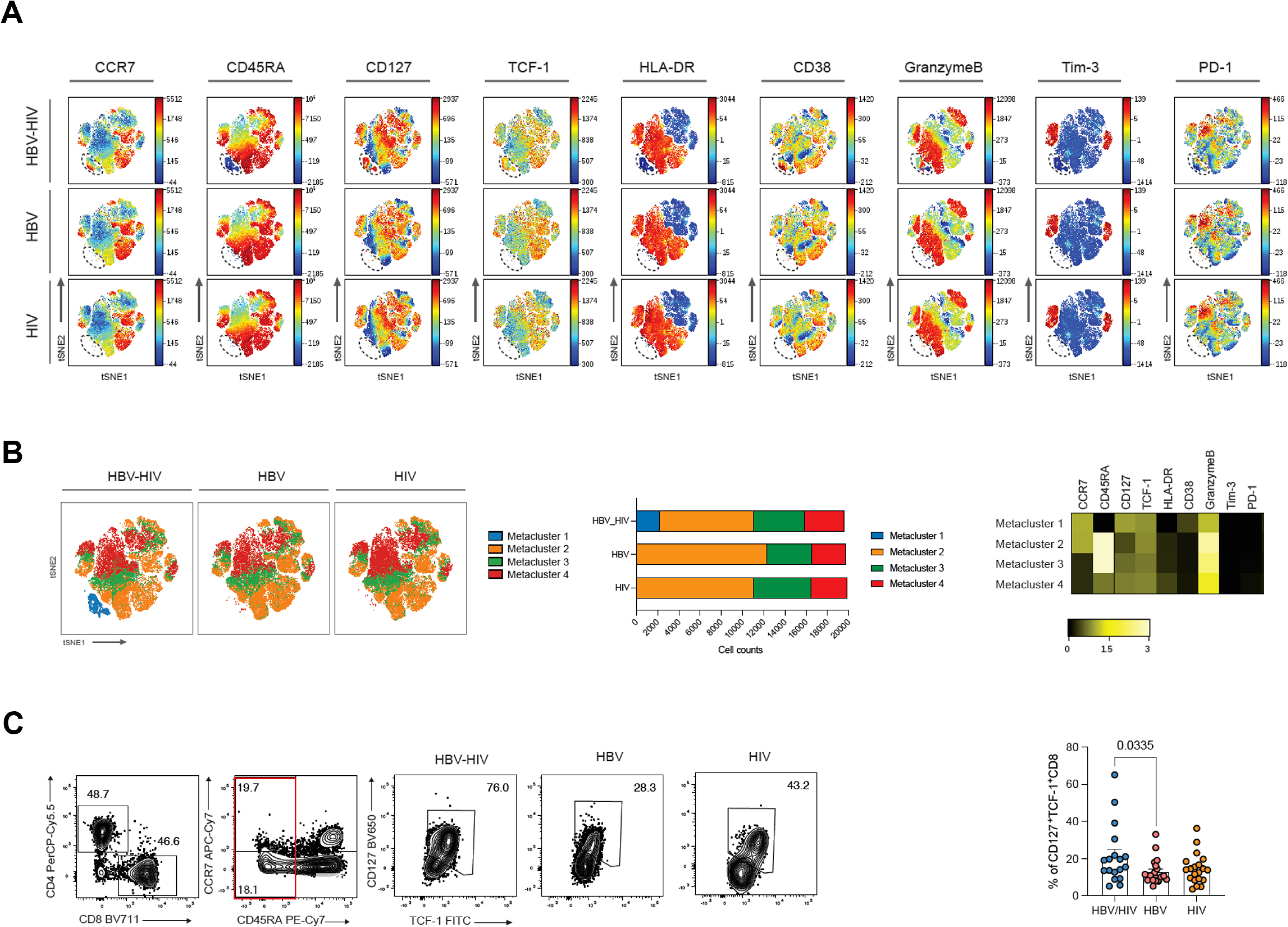
Stem-like CD8+ T cells are enriched in HBV/HIV co-infection. (A) viSNE analysis conducted on CD8+ T cells using concatenated files from HBV/HIV+ (n = 20), HBV+ (n = 20), and HIV+ (n = 21) donors in each study group. Each point on the high-dimensional mapping represents an individual cell, and colour intensity represents the expression of selected markers. (B) viSNE map of FlowSOM metaclusters of CD8+ T cells from people with HBV/HIV, HBV, or HIV. Metaclusters are colour-coded. Cell count of each FlowSOM metaclusters out of total CD8+ T cells (20,000 cells/group). Heatmap of the selected markers for CD8+ T cell clusters. **(C)** Frequency of CD127^+^TCF^-^1^+^CD45RA^+^CCR7^-/low^CD8^+^ T cells via traditional gating in all patient groups. Summary data of the proportion of CD127^+^TCF-1^+^CD45RA^+^CCR^-/low^CD8^+^ T cells in the study groups Significance determined by two-tailed Mann−Whitney U test.

Building on the transcriptomic findings and FlowSOM clustering, we characterised the exhaustion landscape in greater detail across the study groups, with particular attention to co-expression patterns of TCF-1, CD127 and PD-1, signatures associated with Tpex versus terminal exhaustion (Tex) states on global CD8^+^ T cells (**figure 3A-C**). While conventional exhaustion/activation markers (PD-1, HLADR/CD38) showed no significant differences between groups (**supplementary figure 3A-B**), we observed reduced CD127 expression on CD8+ T cells in HBV mono-infection alongside elevated TOX levels (**supplementary figure 3C-D**), robust expression of which results in commitment to Tex^33^. Higher frequencies of CD8^+^ T cells with a stem-cell Tpex phenotype expressing TCF-1^+^CD127^+^PD-1^+^ were identified in people with HBV/HIV compared to either mono-infection group (**figure 3B-C**), mirroring the transcriptomic enrichment of stemness-associated genes in this population. Conversely, Tex CD8+ T cells characterised by TCF-1-CD127-PD-1+ expression were predominant in HBV mono-infection (**figure 3B-C**). While TOX expression was similar across Tpex populations in all groups (**figure 3D**), TCF-1^-^CD127^-^PD-1^+^TOX^+^ cells were significantly increased in HBV mono-infection (**figure 3E**), in keeping with a terminal exhaustion state. Tpex cells resided primarily within the TEM compartment, while Tex cells were enriched in TEMRA populations in HBV mono-infection (**supplementary figure 4A-B**). To further address the differentiation stage of the particular CD127/TFC-1 subsets, we stained for the effector cell molecule granzyme-B. As expected, Tpex populations were further characterised by lower granzyme-B expression^34^; by contrast granzyme-B^+^ Tex CD8^+^ T cells were predominant in HBV mono-infection (**supplementary figure 4C-D**) aligning with the higher cytotoxicity signatures observed in the transcriptomic data. Collectively, our findings reveal distinct CD8+ T cell exhaustion landscapes in HBV versus HBV/HIV co-infection.

**Figure 3.**
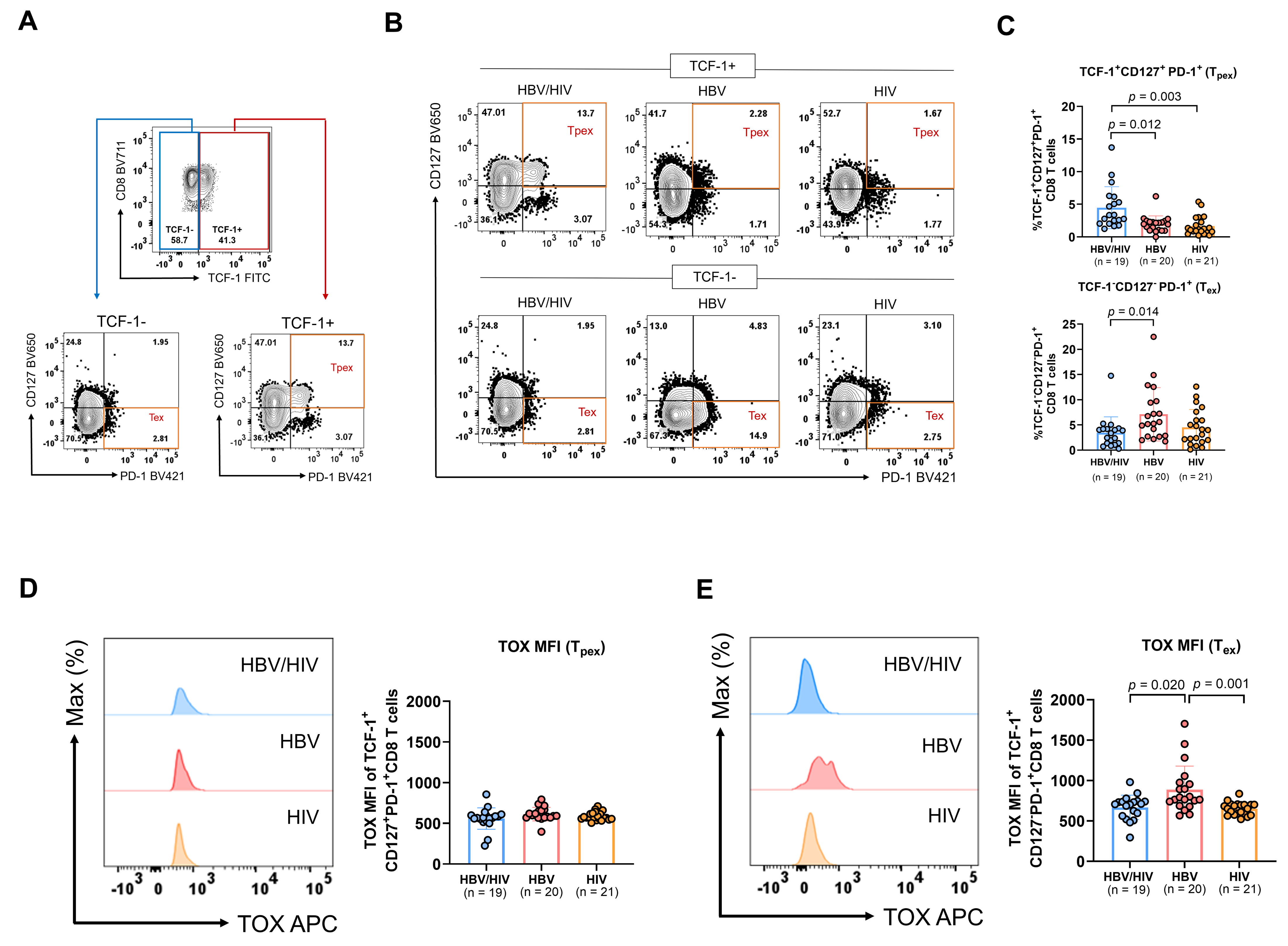
Phenotypic characterisation of Tpex and Tex CD8^+^ T cells in peripheral blood of donors with HBV/HIV, HBV, and HIV infection. (A) Representative flow plots of gating strategy for identifying Tpex and Tex. (B) Representative flow plots of Tpex and Tex profiles identified by expression of TCF-1, CD127 and PD-1 in peripheral blood and (C) summary data from HBV/HIV+ (n = 19), HBV+ (n=20), and HIV+ (n=20) donors. Representative histogram and summary data of TOX MFI on (D) Tpex and (E) Tex CD8^+^ T cells from HBV/HIV+ (n = 19), HBV+ (n=20), and HIV+ (n=20) donors. Bar charts show the median value with interquartile range and each dot represents one donor in a group. Statistical significance was assessed by Kruskal-Wallis with Dunn’s multiple comparison test.

### Higher frequency and polyfunctional HBV-specific CD8^+^ T cell responses in HBV/HIV co-infection compared to HBV

Having established distinct exhaustion profiles in global CD8+ T cell populations, we next sought to determine whether these phenotypic differences translated to functional discrepancies in virus-specific CD8+ T cell responses between individuals with HBV mono-infection and those with HBV/HIV co-infection. Intracellular cytokine staining (ICS) was employed to investigate the composition and polyfunctionality of T cell responses among individuals with HBV/HIV co-infection, as well as those with HBV and HIV mono-infection. PBMCs were stimulated overnight using overlapping peptides targeting the HBV surface (S) and core proteins, alongside CEF and HIV-1 gag peptides within the same individuals.

Notably, after overnight stimulation, higher magnitude HBV-specific responses, identified by co-expression of IFN-γ and TNF-α, were seen in donors with HBV/HIV co-infection relative to donors with HBV (**figure 4A-B**). Over 90% of donors with co-infection demonstrated detectable HBV-specific responses (>0.05% IFN-γ^+^TNF-α^+^ CD8^+^ T cells), compared to 50% in HBV mono-infection (**figure 4C**). Within individual donors, HBV-specific responses were comparable to those against CEF or HIV- 1 gag peptides (**figure 4B**). HIV-1 gag-specific responses were similar between people with HBV/HIV co-infection and those with HIV mono-infection (**figure 4B**). HBV-specific CD8^+^ T cells displayed polyfunctional responses in people with co-infection, particularly against HBV surface protein (**figure 4D**). HBV-specific CD8^+^ T cells primarily displayed EM/TEMRA phenotypes, with core-specific responses in people with co-infection also exhibiting a more notable CM phenotype compared to HBV mono-infection **(Supplemental figure 5A-B**). While people with co-infection showed higher mean frequencies of CEF responses compared to those with mono-infection, these differences were not statistically significant and displayed similar polyfunctional profiles (**figure 4A, B and D**). Despite elevated PD-1 expression on both HBV- and HIV-specific CD8^+^ T cells compared to CEF-specific cells (**supplementary figure 5C-D**), this did not compromise their functional capacity, suggesting PD- 1 expression in this context may not exclusively indicate exhaustion.

**Figure 4.**
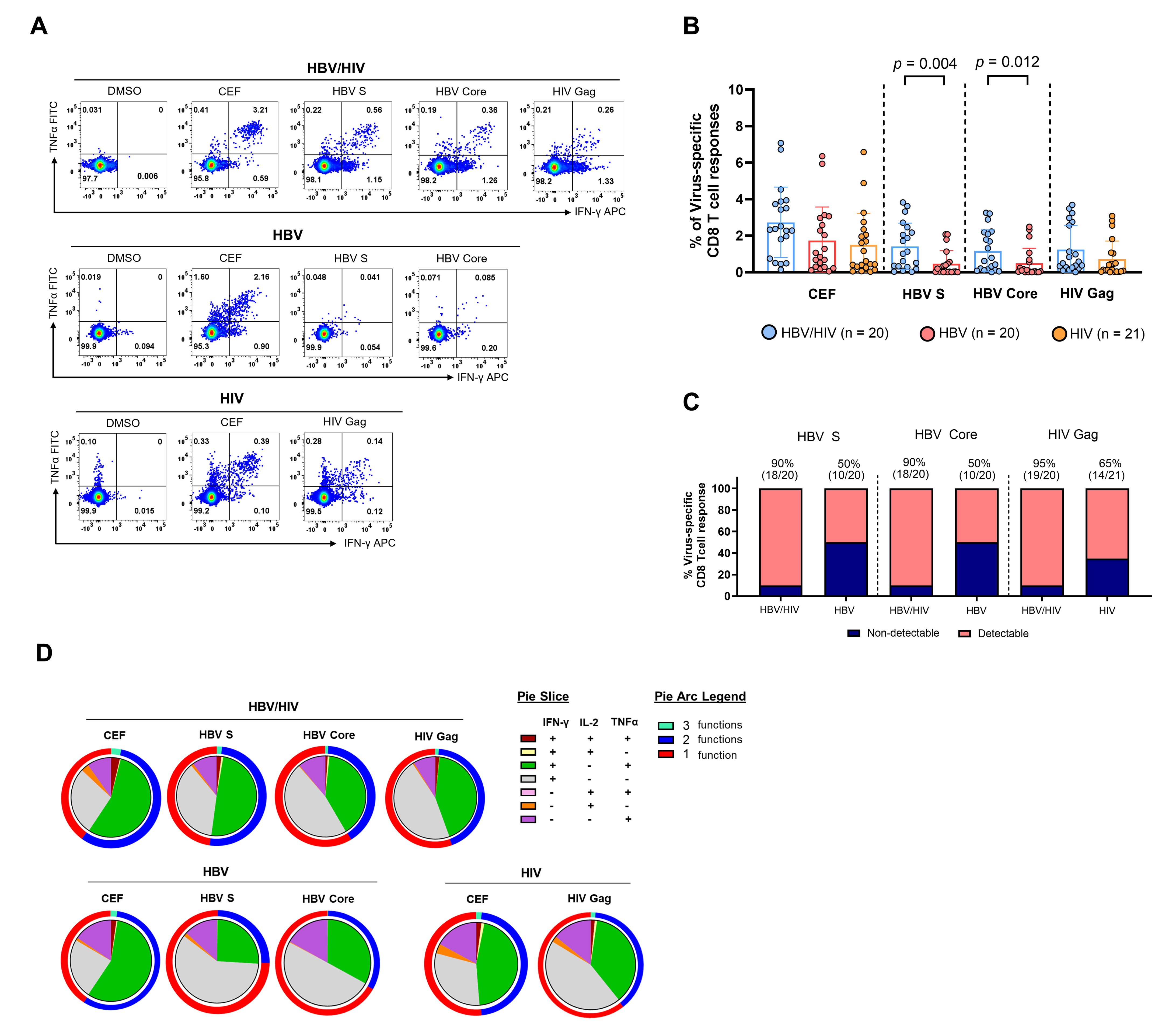
Composition of HBV and HIV-specific CD8+ T cells in HBV/HIV, HBV, and HIV infection. Intracellular cytokine staining (ICS) was performed to examine virus-specific CD8^+^ T cells to the indicated peptide pools in HBV/HIV+ (n = 20), HBV+ (n = 20), and HIV+ (n = 21) donors. (A) Representative flow cytometric plots for the identification of antigen-specific CD8+ T cells based on double expression (IFN-γ^+^TNF-α^+^) following overnight stimulation with DMSO in media (control), CEF pool (positive control) or overlapping HBV-S, HBV-core and HIV-gag peptides. (B) Summary of aggregated CD8+ T-cell responses (IFN-γ^+^TNF-α^+^) against CEF, HBV-S, HBV-core or HIV-gag peptide pools from HBV/HIV+ (n = 20), HBV+ (n = 20), and HIV+ (n = 21) donors. (C) Proportion of detectable and non-detectable virus-specific CD8^+^ T cell responses in HBV/HIV+, HBV+, and HIV+ donors. (D) Pie charts representing the polyfunctional subsets of CEF-, HBV-S-, HBV-core- or HIV-gag-specific CD8^+^ T cell responses with IFN-γ, TNF-α and IL-2 expression, and pie arcs denoting the relative proportions of virus-specific CD8+ T cell response for one(red), two (blue) and three (green) cytokines. Statistical significance was assessed by Kruskal-Wallis with Dunn’s multiple comparison test (*p* < 0.05).

To further characterise HBV-specific CD8^+^ T cells, we employed MHC I dextramers to detect virus-specific CD8^+^ T cells directly *ex vivo* targeting known HLA-A*02-restricted epitopes. We focused on HBV_env_ (combined env_183–191_, env_335–343_, and env _348–357_), pp65_495-504_ and Gag_77-86_ in a subset of HLA- A*02-positive donors with co-infection and detectable responses. We observed that HBV env-specific CD8^+^ T cells displayed higher frequencies of Tpex cells (PD-1^+^/CD127^+^) **(figure 5A)**. Similar trends were observed for both HCMV pp65-specific and Gag-specific CD8^+^ T cells showing enrichment of CD127^+^PD-1^+^ (Tpex phenotype) compared to CD127^-^PD-1^+^ (Tex phenotype) (**supplementary figure 6A, B**). Next, we assessed the expression of the transcription factor TCF-1 and anti-apoptotic molecule BCL-2 that have been shown to best determine the memory-like phenotype and cellular persistence of this subset^35^. CD127^+^PD-1^+^ HBV env-specific CD8^+^ T cells contained higher mean frequencies of TCF-1^+^ and BCL-2^+^ cells compared to CD127^-^PD-1^+^ populations (**figure 5B, C**), with HCMV-specific and HIV Gag-specific cells showing similar patterns(**supplementary figure 6C and D**). These results would be in keeping with the enhanced survival potential of this precursor exhausted subset.

**Figure 5.**
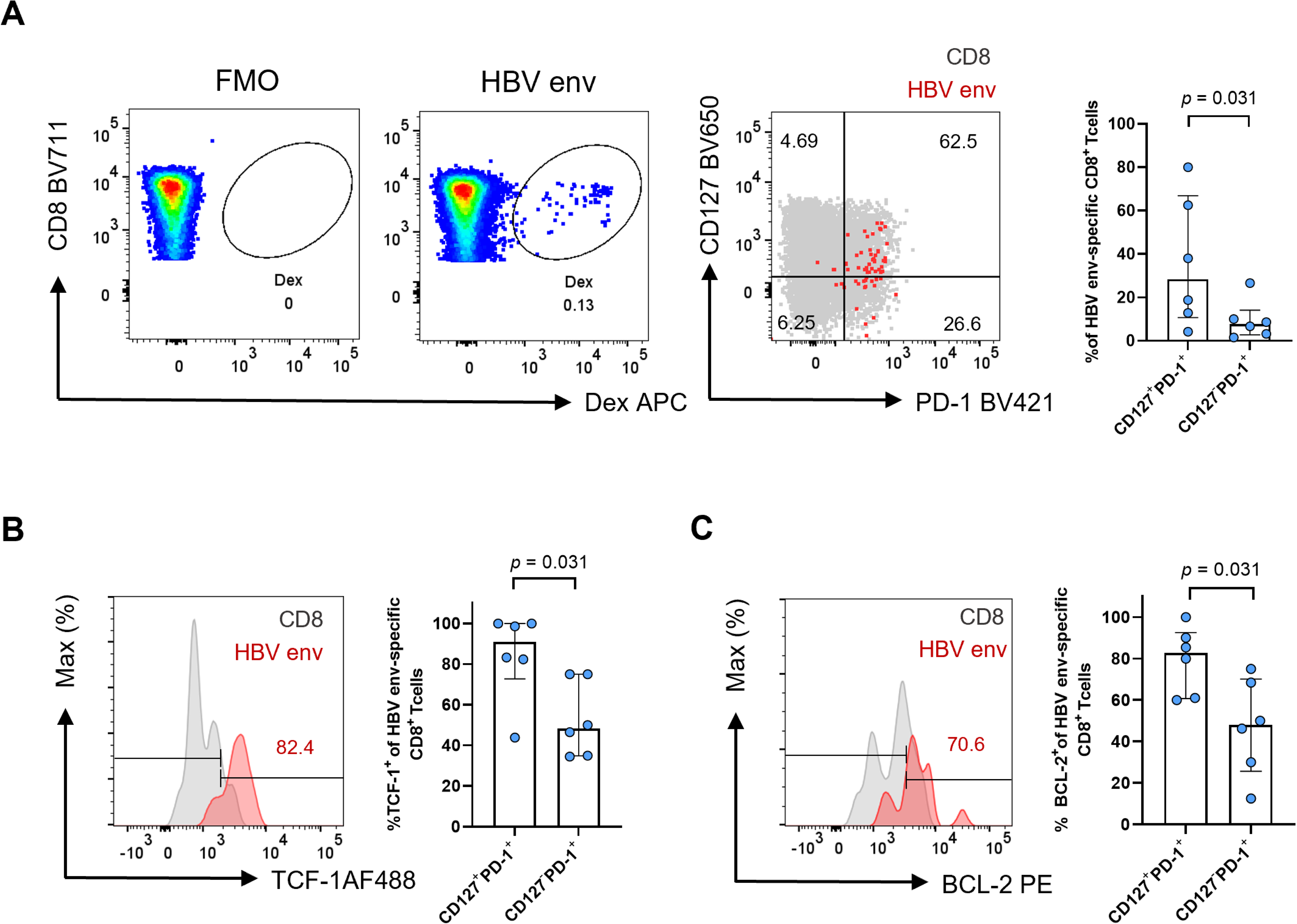
Ex vivo HBV-specific CD8^+^ T cells in HBV/HIV co-infection. (A) a representative plot of HBV_env_–specific CD8^+^ T cell identification by dextramer staining. Representative flow plot (red: HBV_env_–specific CD8^+^ T cell; grey: corresponding global CD8^+^ T cells) and summary data of CD127/PD-1 co-expression analysis of HBV_env_-specific CD8^+^ T cells derived from individuals with HBV/HIV (n = 6). (B) Representative histogram and gating of TCF-1 expression on HBV_env_-specific CD8^+^ T and global CD8^+^ T cells and summary data of TCF-1 expression on HBV_env_-specific CD8^+^ T with respect to CD127/PD-1 subsets. (C) Representative histogram and gating of BCL-2 expression on HBV_env_-specific CD8^+^ T and global CD8^+^ T cells and summary data of BCL-2 expression on HBV_env_-specific CD8^+^ T with respect to CD127/PD-1 subsets. Representative histograms of the individual markers are depicted (grey: corresponding global CD8^+^ T cells and red: HBV epitope-specific CD8^+^ T cells). Bar charts show the median value with interquartile range. Statistics were performed using a two-tailed Wilcoxon matched-pairs signed-rank test (p < 0.05).

### HBsAg levels and treatment duration shape CD8^+^ T cell exhaustion profiles and functional responses

Having established distinct phenotypic and functional differences in CD8^+^ T cell responses between patient groups, we examined potential virological factors that might explain these immunological differences. Quantitative HBsAg analysis revealed significantly lower levels in people with HBV/HIV co-infection compared to those with HBV mono-infection (**figure 6A**), in keeping with our recent observations^14^. A greater percentage of people with HBV/HIV had HBsAg below 50 IU/mL, including some with levels below 1 IU/mL (**figure 6B**), potentially reflecting the longer duration of antiviral therapy in this group (average 14 years versus 4 years in mono-infection, **supplementary table 1**).

**Figure 6.**
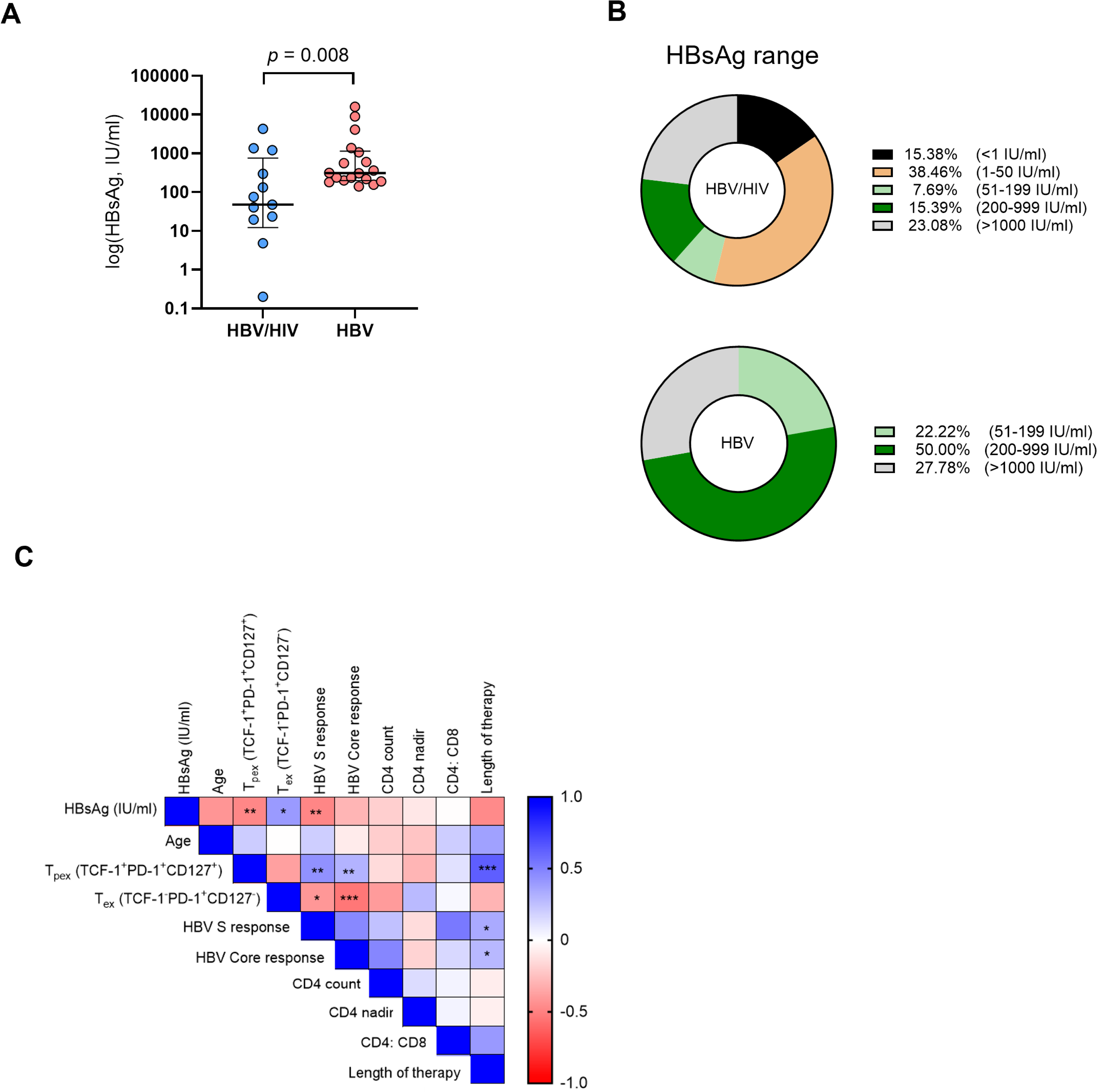
Virological parameters and correlation analysis. (A) HBsAg level quantification (log HBsAg, IU/ml) from individuals with HBV/HIV and HBV. Data shown as scatter plots with the group median and interquartile range indicated. (B) Proportion of HBV/HIV+ or HBV+ donors within a given range of HBsAg level (IU/ml). (C) Correlogram of individuals with HBV/HIV and HBV utilised for phenotypic and functional characterisation. Spearman r values shown from red (−1) to blue (1) with r values indicated by colour. Spearman r value shown. *p<0.05, **p<0.01, and ***p<0.001.

Correlation analyses revealed negative associations between circulating HBsAg levels and both functional HBV-specific responses and Tpex frequencies (**figure 6C**). Conversely, Tpex populations correlated positively with HBV-specific responses and treatment duration, while Tex populations showed inverse relationships with functional responses, particularly against HBV core (**figure 6C**). Treatment duration also positively associated with HBV-specific responses, suggesting progressive immune restoration with sustained viral suppression. We did not detect any associations with age, CD4 count (including CD4 nadir) or CD4:CD8 ratio.

## DISCUSSION

The goal of immune-based strategies for chronic HBV infection is to elicit functional and durable HBV- specific CD8+ T cell responses that can bolster antiviral control. Our findings challenge the conventional understanding of immune responses in HBV/HIV co-infection by demonstrating that individuals with well-controlled HBV/HIV co-infection on long-term suppressive therapy display enhanced preservation of functionally competent HBV-specific CD8^+^ T cells compared to those with HBV mono-infection.

The increased frequency of TCF-1^+^CD127^+^PD-1^+^ (Tpex) CD8^+^ T cells observed in people with co-infection aligns with emerging evidence demonstrating the critical role of these cells in maintaining antiviral responses during human chronic infection^36–40^. These memory-like CD8^+^ T cells, that occur early during infection^41^ ^42^, maintain proliferative capacity despite chronic antigen exposure and provide the proliferative burst following PD-1 pathway blockade^16^ ^17^ ^31^ ^34^. Our findings extend this concept to HBV/HIV co-infection, where we observe higher frequencies of Tpex cells and more robust HBV- specific responses than in HBV mono-infection. The preservation of a stem-like CD8^+^ T cell pool capable of sustained antiviral responses may facilitate HBsAg clearance in people with co-infection, who demonstrate higher rates of functional cure^12^ ^13^. Furthermore, these results align with our recent work demonstrating enhanced NK cell ADCC in co-infection^14^, suggesting a better-preserved immunological profile favouring viral control.

The transcriptional signature of CD8^+^ T cells in HBV/HIV co-infection, suggests adaptive mechanisms allowing CD8^+^ T cells to navigate a potentially more complex immunological landscape due to the presence of two chronic viral infections. Upregulation of TCR signalling and inhibitory pathway genes (e.g., *CD160*, *TIGIT*, *PDCD1*) alongside genes involved in inflammation regulation (*CHI3L2*, *KLF10*) and mitochondrial function (*CMC1*) suggests a balanced immune response that prevents excessive inflammation while maintaining functional responses. Along these lines, PD-1 expression on Tpex virus-specific CD8^+^ T cells likely shields these populations from excessive TCR stimulation, preserving their stem-like properties^15^. This interpretation is supported by our observation that these populations maintain high expression of BCL-2 and TCF-1, markers associated with memory potential and cellular longevity, despite chronic antigenic stimulation^39^. In contrast, HBV mono-infection exhibited a profile skewed toward terminal exhaustion, with elevated TOX expression, that transcriptionally/epigenetically programmes CD8^+^ T cell exhaustion^33^, and linked to CD8^+^ T cell dysfunction during cHBV^43^.

Our findings echo recent observations demonstrating that exhaustion profiles in global CD8+ T cells can predict functional capacity of HBV-specific responses^6^. The apparent paradox of better T cell functionality in co-infection could be explained by several factors. Significantly lower HBsAg levels observed in people with co-infection likely reduce antigen-driven exhaustion, supported by the negative correlation between HBsAg and functional T cell responses. This aligns with findings from previous studies demonstrating the detrimental effects of sustained HBsAg exposure on virus-specific T cells^8^. Earlier initiation of potent HBV-active therapy in people diagnosed with HIV leads to more rapid suppression of viral replication, as reflected by the longer average treatment duration in our co-infected cohort. This early intervention likely preserves the Tpex pool before terminal exhaustion occurs, consistent with our observation that treatment duration positively correlates with functional responses and Tpex frequencies. Additionally, the immunological milieu created by HIV infection^44^ may modulate HBV-specific responses through altered cytokine profiles or reduced immune regulatory mechanisms. Our recent findings of enhanced proportions of adaptive/memory like NK cells in HBV/HIV co-infection^14^, with less inherent potential for excessive negative regulation of T cell responses^45^ ^46^, supports this broader immune remodelling hypothesis.

Multiple studies have demonstrated that targeting the PD-1 pathway can enhance HBV-specific CD8^+^ T cell function^47^, and affect the HIV viral reservoir and virus-specific responses^48^ ^49^. Our data suggest that such approaches may be particularly effective in people with co-infection who maintain a higher proportion of Tpex cells. Similarly, approaches to target the TCF-1 pathway^50^ and enhance stemness through IL-15 signalling^51^ or TOX manipulation even after Tex establishment^52^ could therefore provide therapeutic opportunities to rewire Tex cells in chronic infections.

Several limitations of our study warrant consideration. As a primarily cross-sectional investigation conducted on peripheral blood, we cannot directly extrapolate our findings to hepatic tissue. The observed differences may be significantly influenced by our cohort characteristics, particularly regarding infection routes and treatment histories, which may limit the generalisability of our findings. In Western settings, HBV/HIV co-infection typically occurs through sexual transmission in adulthood, while HBV mono-infection often results from vertical transmission, leading to fundamental differences in immune priming and tolerance. Additionally, our analysis focused on surface and core HBV-specific responses; extending this to other antigens would provide a more comprehensive understanding of virus-specific responses, as exhaustion profiles differ based on targeted antigens^6^ ^39^.

Future longitudinal studies should examine how these immunological differences evolve over time in response to therapy and correlate with clinical outcomes. Tissue-based analyses would provide insight into the hepatic immune microenvironment, while functional studies examining the response of Tpex cells to immunotherapeutic interventions could inform personalised treatment approaches based on exhaustion phenotypes rather than infection status alone. Additional work beyond the scope of this study, will aim to provide more mechanistic insights into the maintenance of Tpex populations in co-infection that could identify novel targets for enhancing immune function.

Despite these limitations, our study provides the first evidence that in individuals with well-controlled HBV/HIV co-infection in the current ART era, CD8^+^ T cell responses are more robust and retain stem-like properties that support ongoing antiviral function. These findings underscore the benefits of early antiretroviral intervention and highlight TCF-1-expressing stem-like CD8^+^ T cells as a promising target for immunotherapeutic strategies aimed at achieving functional cure in chronic HBV infection.

## Data Availability

All data relevant to the study are included in the article or uploaded as online supplemental information.
Raw data for single cell sequencing reported in this study have been deposited in Gene Expression Omnibus (GSE241183).

## Contributors

JP performed experiments, acquisition of data, analysis, and drafting of the manuscript; BS, AA, EAB, JK, FG, SP, KDC, NFP, NH, SK, performed experiments and contributed to data acquisition and analysis. IG, FB, SK, PS, SB, PTFK, MKM, RBS and USG contributed clinical samples, data interpretation and critical editing of the manuscript. DP contributed to the conception and design of the study, data interpretation, critical revision of the manuscript, and study supervision.

## Data availability statement

All data relevant to the study are included in the article or uploaded as online supplemental information.

Raw data for single cell sequencing reported in this study have been deposited in Gene Expression Omnibus (GSE241183).

## Ethics statements

### Patient consent for publication

Not applicable. Patients or the public were not involved in the design, or conduct, or reporting, or dissemination plans of our research

### Ethics approval

The study was approved by the local ethics committee (Berkshire (REC 16/SC/0265) and London Bridge (REC 17/LO/0266) and conformed to the Helsinki declaration principles.

